# Short-term outcomes of asphyxiated neonates depending on outborn versus inborn status

**DOI:** 10.1101/2024.03.05.24303778

**Authors:** Nora Bruns, Nadia Feddahi, Rayan Hojeij, Rainer Rossi, Christian Dohna-Schwake, Anja Stein, Susann Kobus, Andreas Stang, Bernd Kowall, Ursula Felderhoff-Müser

## Abstract

**Importance:** In neonates with birth asphyxia (BA) and hypoxic ischemic encephalopathy, therapeutic hypothermia (TH), initiated within six hours, is the only safe and established neuroprotective measure to prevent secondary brain injury. Infants born outside of TH centers have delayed access to cooling.

**Objective:** To compare in-hospital lethality, occurrence of seizures, and functional status at discharge in newborns with BA depending on postnatal transfer to another hospital within 24 hours of admission (outborn versus inborn).

**Design:** Nationwide retrospective cohort study from a comprehensive hospital dataset using codes of the International Classification of Diseases, 10^th^ modification (ICD-10). Clinical and outcome information was retrieved from diagnostic and procedural codes. Hierarchical multilevel logistic regression modelling was performed to quantify the effect of being outborn on target outcomes.

**Setting:** All admissions to German hospitals 2016 – 2021.

**Participants:** Full term neonates with birth asphyxia (ICD-10 code: P21) admitted to a pediatric department on their first day of life.

**Exposures:** Transfer to a pediatric department within 24 hours of admission to an external hospital (=outborn).

**Main outcomes:** In-hospital death; secondary outcomes: seizures and pediatric complex chronic conditions category (PCCC) ≥ 2.

**Results:** Of 11,703,800 pediatric cases, 25,914 fulfilled the inclusion criteria. Outborns had higher proportions of organ dysfunction, TH, organ replacement therapies, and neurological sequelae in spite of slightly lower proportions of maternal risk factors. The adjusted odds ratios (OR) for death, seizures, and PCCC ≥ 2 were 4.08 ((95 % confidence interval 3.41 – 4.89), 2.99 (2.65 – 3.38), and 1.76 (1.52 – 2.05), respectively, if infants were outborn compared to inborn. A subgroup analysis among infants receiving TH (n = 3,283) found less pronounced adjusted ORs for death (1.67 (1.29 – 2.17)) and seizures (1.26 (1.07 – 1.48)) and inversed effects for PCCC ≥ 2 (0.81 (0.64 – 1.02)).

**Conclusion and relevance:** This comprehensive nationwide study found increased odds for adverse outcomes in neonates with BA who were transferred to another facility within 24 hours of hospital admission. Obstetrical units should be linked to a pediatric department to minimize risks of postnatal emergency transfer. Collaboration and coordination between centers should be improved to balance geographical coverage of different level care facilities.

**Key points:** *Question:* How does outcome in neonates with birth asphyxia differ depending on postnatal transfer status to a pediatric department?

*Findings:* In this comprehensive nationwide cohort study from administrative data including 35,250 cases, outborns had increased odds for death, seizures, and impaired functioning in spite of similarly distributed maternal risk factors and lower prevalence of infant risk factors.

*Meaning:* To prevent postnatal transfer and potential delays in optimal neonatal care, maternity hospitals should invariably be combined with a pediatric unit. Neonatal emergency trainings and telemedicine may help to attenuate the adverse effects of being born in a non-TH center and in settings without access to a pediatric unit.

## Introduction

Lack of oxygen around birth (perinatal asphyxia) leads to impaired metabolic supply of vital organs and may cause severe and potentially irreversible damage, particularly to the brain. The resulting complication hypoxic-ischemic encephalopathy (HIE) is one of the most common causes of perinatal morbidity and mortality and a major contributor to long-term neurological impairment. In Western societies, up to three out of 1000 live births are affected from moderate or severe HIE, which is associated with high individual burden and relevant costs for healthcare systems [1-3].

Therapeutic hypothermia (TH) as the only established and safe neuroprotective measure to reduce case-fatality and secondary neurological damage should be initiated as fast as possible, but no later than 6 hours following the insult [4, 5]. Diagnostic criteria used by neonatal resuscitation teams to initiate TH for moderate and severe HIE have been widely adopted from large international multicenter trials and are part of national guidelines [4-7]. They include early identification of clinical risk factors and neurological scores, preferably supported by amplitude-integrated electroencephalography for risk stratification, early seizure detection, and prediction of neurological outcome [8, 9]. Evidence from both animal and human studies suggest that early achievement of the target temperature of 33°-34 °C is beneficial for the neurological outcome. Servo-controlled devices provide optimal results when administering targeted hypothermia. However, hypothermia can be applied passively or via non-servo-controlled devices as a bridging technology during transport or if servo-control is unavailable. It has been reported that newborns with HIE born outside of hospitals with a Neonatal Intensive Care Unit (NICU) reach the target cooling temperature later and develop poorer outcomes compared to inborns [10-12].

For Germany, it is unknown how lethality and short-term survival following BA and HIE vary depending on postnatal transfer. The aim of this study was to compare in-hospital lethality and short-term outcomes according to inborn versus outborn birth status in neonates with BA. For this purpose, we conducted a large retrospective cohort study of neonates with BA in a nationwide comprehensive dataset across Germany.

## Methods

The German hospital dataset (GHD) is a nationwide dataset comprising all hospitalizations in public hospitals in the country. Since 2004, German hospitals have been receiving reimbursement based on diagnosis related groups (DRG). As per §21 KHEntgG, it is mandated by law that German hospitals share data on all hospital admissions with the Hospital Remuneration System (InEK). Once subjected to plausibility checks, this data is anonymized and then sent to the Federal Statistical Office (FSO). Given that the provision of hospitalization data is obligatory for reimbursement, hospitals are strongly motivated to provide comprehensive data.

A retrospective cohort study using secondary data on admissions to all hospitals in Germany between 2016 and 2021 was conducted in the GHD. The dataset provided by the FSO for access at the research data center Düsseldorf (Germany) was pre-filtered for cases < 18 years of age at admission. Detailed information regarding the structure of the GHD can be obtained from the FSO, and further details on the process of data access are available at https://www.forschungsdatenzentrum.de/en/health/drg.

### Analyzed cases

Eligible cases were identified via the International Classification of Diseases, 10th Edition, German Modification (ICD-10-GM). Cases with birth asphyxia (BA) as primary or secondary discharge diagnosis (ICD code: P21) were eligible. The admission reason had to be “in-hospital birth” or “referral from an external hospital within 24 hours of admission to the external center” and admission age had to be ≤ 2 days. Cases born in maternity hospitals without a pediatric department were analyzed if they died before transfer on the first day of life.

### Data extraction

Secondary diagnoses and medical procedures were extracted via ICD codes and codes for surgeries and procedures (Operations-und Prozedurenschlüssel, OPS) to assess the course of disease, interventions, organ failure, clinical outcomes, and maternal and infant risk factors. Organ dysfunction was assessed by extracting binary scores indicating organ dysfunction and summed up (neonatal organ failure (NOF) score). Functional outcome was assessed using the Pediatric Complex Chronic Conditions (PCCC) Classification with modifications due to lack of information on device prescription in the GHD [13]. The PCCC was designed to identify and quantify conditions in children that are likely to persist for at least one year. Congenital malformations are extensively considered in the PCCC calculation. All extracted and newly calculated items are listed in supplementary table 1 and the items used to calculate the PCCC is detailed in supplementary table 2.

### Primary and secondary outcomes

The unit of analysis was hospital admission for/with BA. The primary outcome was in-hospital death. Secondary endpoints were seizures and a PCCC score ≥ 2.

### Center description

Center characteristics were empirically derived from the complete pediatric dataset (11,703,800 cases) using the institutional identifier. Children’s hospitals were defined as institutions that used pediatric department (PD) codes during the study period. Cooling centers were defined as institutions that conducted TH in neonates with HIE during the study period.

### Definition of outborn versus inborn cases

Only cases treated in pediatric wards were analyzed. Cases were considered inborn if the reason for hospital admission was “in-hospital birth”. Cases were considered outborn if the reason for admission was “referral from an external hospital within 24 hours of admission to the external hospital” or if they died in a hospital without PD.

### Out-of-hospital births

Due to the admission criteria “in-hospital birth” or “referral from external hospital”, infants born outside of hospitals were excluded. A marginal possibility that an infant was born out of hospital and then transferred from the initial hospital within 24 hours probably applies only for a minor fraction of cases.

### Missing Data

No data were missing for age and primary diagnoses. Missing secondary diagnoses and procedures could not be detected due to impossible differentiation between cases with truly absent versus uncoded diagnoses. We assumed that diagnoses and procedures that are well-reimbursed were coded comprehensively, and therefore centered data extraction on these specific codes.

### Statistical analyses

Continuous variables are presented as mean for symmetrical and unimodal distributions and as median for skewed distributions. Discrete variables are summarized as counts and relative frequencies.

Minimally sufficient adjustment sets for regression analyses were identified via causal diagrams based on the theory of directed acyclic graphs (DAG) [14, 15] as recommended for empirical pediatric and critical care research [16, 17] (Supplementary figure 1). Because two equally plausible causal diagrams were identified that affected the minimally sufficient adjustment sets, a sensitivity analysis was conducted. The basic adjustment set was built on the assumption that the birth hospital setting (and thus outborn/inborn status) determines the severity of asphyxia and comprised the birth weight (in grams), maternal risk factors, and congenital malformations. The alternative adjustment set for sensitivity analyses was based on the assumption that the severity of asphyxia causes the outborn/inborn status to change because severe asphyxia prompts transfer to a high-level center. This alternative adjustment set additionally contained the severity of asphyxia as extracted from ICD-10 codes. The birth hospital was also part of the adjustment set but was unobserved in outborns. Because hospitals frequently transfer their patients to collaborating hospitals and because patients cared for in the same center were not independent, we accounted for clustering (correlation) within centers by hierarchical modeling with the treating (discharging) hospital (identified via the institutional identifier) as random effect in all analyses [18, 19].

Logistic regression was performed for the primary and secondary endpoints. A subgroup analysis including only infants who received TH was additionally conducted using the basic and the alternative adjustment sets described above.

### Software

All calculations were carried out using SAS release 9.4 and SAS Enterprise Guide 7.1 (SAS Institute, Cary, North Carolina, USA).

## Results

Of 11,703,800 pediatric cases discharged between 1^st^ January 2016 and 31^st^ December 2021, 69,033 were neonatal cases with BA admitted to hospital on the first day of life. 33,783 cases were excluded because they were not admitted to pediatric departments or remained in the external hospital for > 24 hours. Of the remaining 35,250 cases, 9,336 were excluded because of preterm birth, leaving 25,914 cases that were eligible for analysis (Figure 1). In total, 3,290 (12.9 %) infants received TH and 755 (3.0 %) infants died. Of these, 62 (8.2 %) infants died in hospitals without PD. Outborns were more often treated in cooling centers than inborns and had a higher proportion of severe asphyxia (Table 1). Maternal risk factors were slightly higher in inborns, as were child-associated risk factors such as hypo- or hypertrophy. Congenital malformations were similarly distributed between groups (Table 1). Organ failure, neurological complications, application of TH and need for extracorporeal organ support (ECMO, Dialysis) were coded more frequently in outborns (Table 2). Time from hospital admission to death was longer in outborns between both groups, with higher lethality among outborns (6.3 % vs. 2.1 %). In survivors, the median length of hospital stay was slightly shorter and median discharge PCCC scores were lower (Table 2). The composite outcome death or PCCC ≥ 2 was observed in 9.0 % of inborns versus 14.7 % of outborns.

**Table 1:**
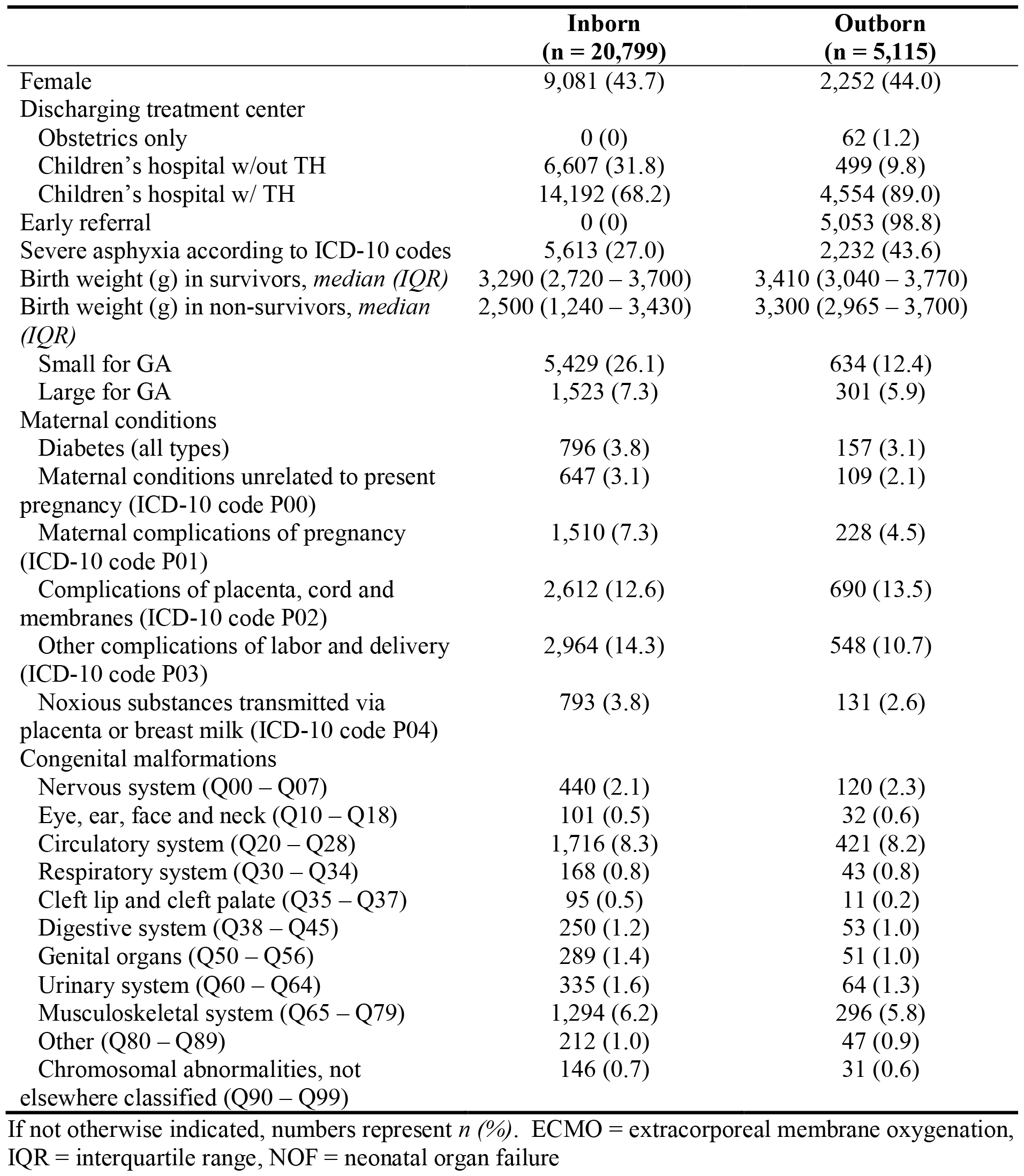
Baseline characteristics of full term neoantes with birth asphyxia treated in pediatric departments in Germany, 2016 – 2021

|  | **Inborn**<br>**(n = 20,799)** | **Outborn**<br>**(n = 5,115)** |
| --- | --- | --- |
| Female | 9,081 (43.7) | 2,252 (44.0) |
| Discharging treatment center |  |  |
| Obstetrics only | 0 (0) | 62 (1.2) |
| Children's hospital w/out TH | 6,607 (31.8) | 499 (9.8) |
| Children's hospital w/ TH | 14,192 (68.2) | 4,554 (89.0) |
| Early referral | 0 (0) | 5,053 (98.8) |
| Severe asphyxia according to ICD-10 codes | 5,613 (27.0) | 2,232 (43.6) |
| Birth weight (g) in survivors, *median (IQR)* | 3,290 (2,720 – 3,700) | 3,410 (3,040 – 3,770) |
| Birth weight (g) in non-survivors, *median (IQR)* | 2,500 (1,240 – 3,430) | 3,300 (2,965 – 3,700) |
| Small for GA | 5,429 (26.1) | 634 (12.4) |
| Large for GA | 1,523 (7.3) | 301 (5.9) |
| Maternal conditions |  |  |
| Diabetes (all types) | 796 (3.8) | 157 (3.1) |
| Maternal conditions unrelated to present pregnancy (ICD-10 code P00) | 647 (3.1) | 109 (2.1) |
| Maternal complications of pregnancy (ICD-10 code P01) | 1,510 (7.3) | 228 (4.5) |
| Complications of placenta, cord and membranes (ICD-10 code P02) | 2,612 (12.6) | 690 (13.5) |
| Other complications of labor and delivery (ICD-10 code P03) | 2,964 (14.3) | 548 (10.7) |
| Noxious substances transmitted via placenta or breast milk (ICD-10 code P04) | 793 (3.8) | 131 (2.6) |
| Congenital malformations |  |  |
| Nervous system (Q00 – Q07) | 440 (2.1) | 120 (2.3) |
| Eye, ear, face and neck (Q10 – Q18) | 101 (0.5) | 32 (0.6) |
| Circulatory system (Q20 – Q28) | 1,716 (8.3) | 421 (8.2) |
| Respiratory system (Q30 – Q34) | 168 (0.8) | 43 (0.8) |
| Cleft lip and cleft palate (Q35 – Q37) | 95 (0.5) | 11 (0.2) |
| Digestive system (Q38 – Q45) | 250 (1.2) | 53 (1.0) |
| Genital organs (Q50 – Q56) | 289 (1.4) | 51 (1.0) |
| Urinary system (Q60 – Q64) | 335 (1.6) | 64 (1.3) |
| Musculoskeletal system (Q65 – Q79) | 1,294 (6.2) | 296 (5.8) |
| Other (Q80 – Q89) | 212 (1.0) | 47 (0.9) |
| Chromosomal abnormalities, not elsewhere classified (Q90 – Q99) | 146 (0.7) | 31 (0.6) |

If not otherwise indicated, numbers represent *n (%)*. ECMO = extracorporeal membrane oxygenation, IQR = interquartile range, NOF = neonatal organ failure

**Table 2:** Neonatal outcomes of full term neonates with birth asphyxia treated in pediatric departments in Germany, 2016 – 2021

|  | Inborn<br>(n = 20,799) | Outborn<br>(n = 5,115) |
| --- | --- | --- |
| Birth injuries |  |  |
| Central nervous system | 139 (0.7) | 56 (1.1) |
| Peripheral nervous system | 200 (1.0) | 51 (1.0) |
| Skeletal | 224 (0.8) | 56 (0.9) |
| Other | 255 (1.2) | 46 (0.9) |
| Organ failure |  |  |
| NOF score survivors, *median (IQR)* | 0 (0 – 1) | 1 (0 – 1) |
| NOF score non-survivors, *median (IQR)* | 2 (1 – 3) | 2 (1 – 3) |
| Cardiovascular | 3,014 (14.5) | 1,046 (20.4) |
| Pulmonary | 9,535 (45.8) | 2,611 (51.0) |
| Coagulation | 1,573 (7.6) | 1,042 (20.3) |
| Renal | 157 (0.8) | 98 (1.9) |
| Hepatic | 44 (0.2) | 29 (0.6) |
| Organ support |  |  |
| Mechanical ventilation | 10,494 (50.5) | 2,867 (56.1) |
| Duration of mechanical ventilation in survivors, (hours) *median (IQR)* | 22 (5 – 111) | 82 (11 – 139) |
| Dialysis | 12 (0.06) | 7 (0.14) |
| Extracorporeal membrane oxygenation | 10 (0.05) | 17 (0.33) |
| Therapeutic hypothermia | 1,853 (8.9) | 1,437 (28.0) |
| Neurologic complications |  |  |
| Brain edema | 89 (0.4) | 77 (1.5) |
| Coma | 141 (0.7) | 140 (2.7) |
| Seizures or epileptic state | 971 (4.7) | 699 (13.7) |
| Epileptic state | 27 (0.1) | 14 (0.2) |
| Hypoxic ischemic encephalopathy | 971 (4.7) | 757 (14.8) |
| Hydrocephalus | 57 (0.3) | 12 (0.2) |
| Deceased | 434 (2.1) | 321 (6.3) |
| PCCC $\geq 2$ in survivors | 1,595 (7.7) | 507 (9.9) |
| Composite outcome death or PCCC $> 2$ | 1,863 (9.0) | 753 (14.7) |
| Pediatric complex chronic conditions classification in survivors, *median (IQR)* | 0 (0 – 1) | 1 (0 – 1) |
| Length of hospital stay in survivors (days), *median (IQR)* | 5 (4 – 11) | 6 (3 – 12) |
| Time from admission to death (days), *median (IQR)* | 3 (1 – 10) | 1 (1 – 6) |

All numbers represent *n (%)* unless otherwise indicated. NOF = Neonatal organ failure, PCCC = Pediatric complex chronic conditions classification

**Figure 1:**
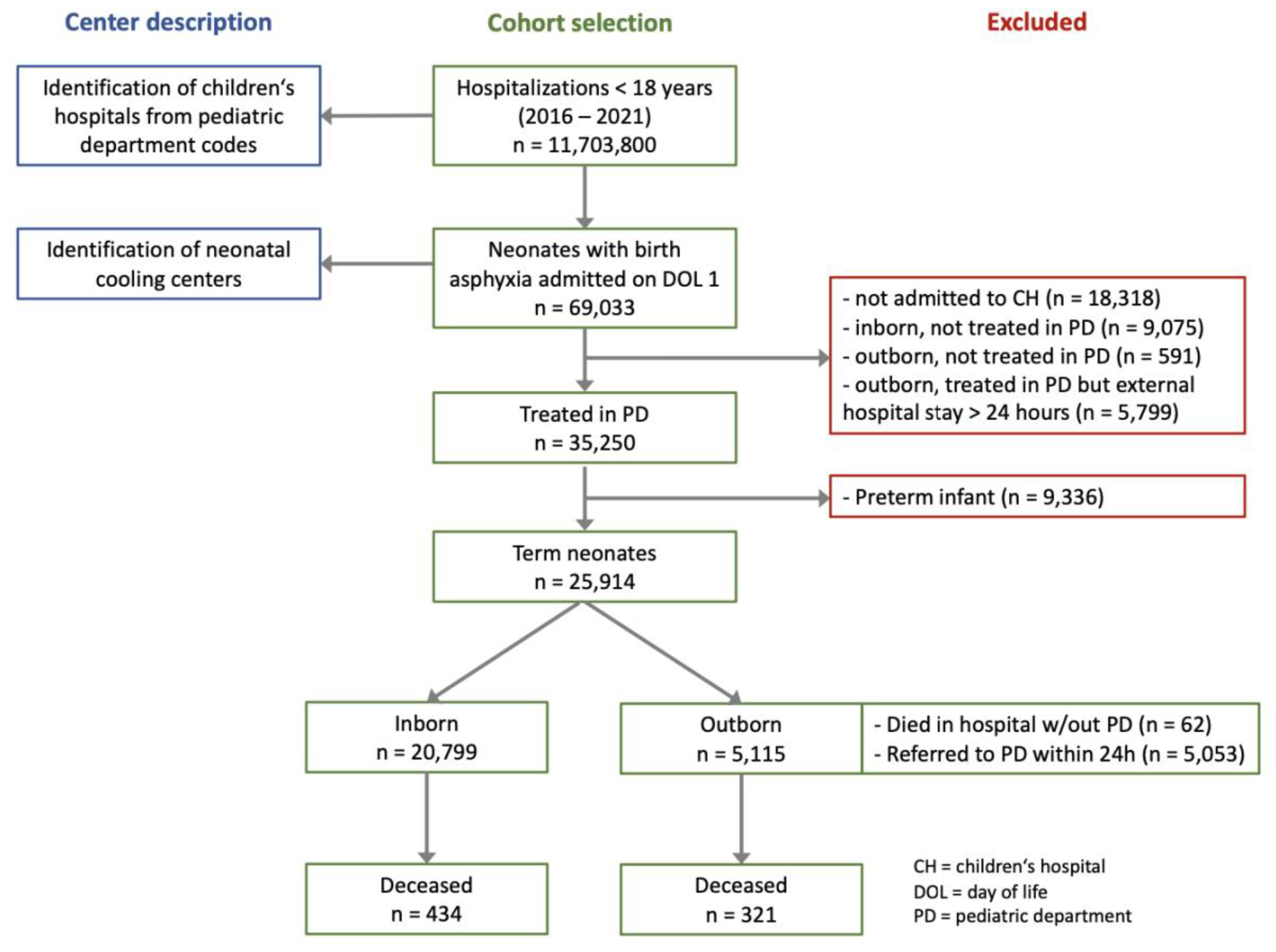
Flow chart of case selection and derivation of center description for inborn and outborn full term neonates with birth asphyxia in Germany, 2016 – 2021

Hierarchical multilevel regression analyses of the overall cohort yielded increased odds ratios (OR) for death (OR 4.08 (95 % CI 3.41 – 4.89)), seizures (2.99 (2.65 – 3.38)), PCCC ≥ 2 (OR 1.76 (1.52 – 2.05)) (Figure 2) in outborns compared to inborns. Results of the sensitivity analysis including adjustment for severity of asphyxia pointed in the same direction (death: 3.00 (2.50 – 3.59), seizures 2.37 (2.09 – 2.69), PCCC ≥ 2 1.39 (1.19 – 1.62).

**Figure 2:**
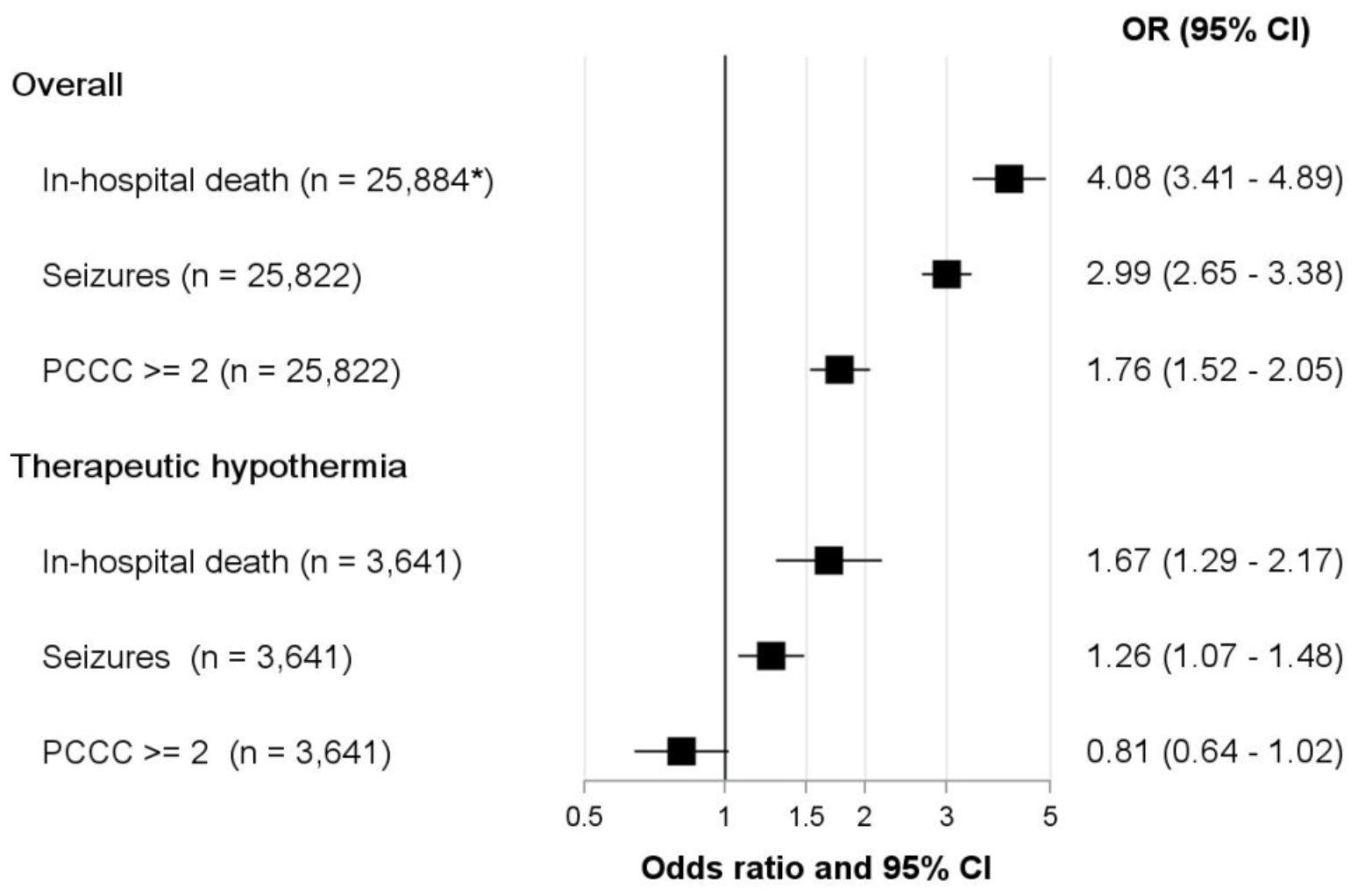
Adjusted odds ratios for death, seizures, and pediatric complex chronic conditions category ≥ 2 in outborn versus inborn neonates with birth asphyxia in Germany, 2016 – 2021. *including 62 neonates that died in hospitals without pediatric department

Results of the subgroup analyses of 3,290 cases receiving TH were less pronounced but pointed in the same direction with an OR for death of 1.67 (1.29 – 2.17) and 1.26 (1.07 – 1.48) for seizures in outborns. However, the OR for PCCC ≥ 2 was 0.81 (0.64 – 1.02) in outborns (Figure 2). The sensitivity analysis including adjustment for severity of asphyxia showed an OR for death of 1.63 (1.26 – 2.12), seizures 1.23 (1.05 – 1.45), and PCCC ≥ 2 0.79 (0.63 – 1.00).

## Discussion

This study, based on the comprehensive nationwide German hospital database, investigated the impact of being born in a maternity hospital without direct access and consequently the need für postnatal transport to a pediatric department with appropriate level of care (outborn) on lethality and short-term outcomes in newborns with birth asphyxia. We found higher lethality and poorer functional outcomes in outborns compared to inborns after control for confounding. These findings were less pronounced in the subgroup of infants who received therapeutic hypothermia with respect to death and seizures. For infants receiving hypothermia that survived PCCC ≥ 2 was more frequent in the inborn group. Outborns receiving TH after BA suffered more frequently from multiorgan failure, suggesting that they were referred either for cooling itself or for management of severe organ dysfunction.

The findings of this study suggest that being born in a setting unprepared to treat the encountered severity of asphyxia and/or associated organ dysfunction is associated with higher lethality and poorer short-term outcomes. During the support of transition in newborns and newborn resuscitation, small differences in management add up and can accumulate to large differences downstream. Thus, the turning point from deterioration to stabilization is not predetermined by child-inherent factors alone, but also depends on the type and effectiveness of measures immediately after birth. In the setting of an asphyxiated infant born outside the reach of a neonatal resuscitation team, potential contributing factors include suboptimal resuscitation by non-neonatal staff, delayed achievement of physiological and metabolic homeostasis after the initial stabilization, lack of diagnostic tools to determine the severity of HIE, delayed transport to a neonatal care center, and the transport itself [10, 20, 21]. These factors are inseparable from the effects of delayed initiation of TH as the only effective measure for neuroprotection following HIE. Above that, the need for TH partially depends on the initial postpartum management.

It is well-acknowledged that TH has to be initiated within six hours after birth and that a delay in the onset of cooling worsens outcomes (“time is brain”) [22, 23]. Outborn infants frequently do not have access to cooling within the critical time window and are either not offered TH or reach the target temperature later [11, 24, 25]. Strategies to overcome the delayed TH onset by active or passive cooling prior to arrival at the cooling center require training and bring along reduced efficiency and risk of overcooling [26].

To further investigate the most severely affected infants – the subgroup receiving TH – we conducted a sensitivity analysis. According to our causal diagram, the application of TH is a mediator of the investigated outcomes, as performed in the most severely affected infants who are at highest risk for brain injury and adverse outcomes. Even though controlling for a mediator will attenuate the observed effects [27, 28], we still observed higher odds ratios for lethality and the occurrence of seizures in outborns in this subanalysis. This suggests that the effect of being outborn is not fully compensated by the application of TH. The effect reversal for PCCC scores ≥ 2 in surviving outborns receiving TH can potentially be explained by the higher lethality in this group, causing depletion of susceptible infants. Unfortunately, we were not able to measure or characterize the additional factors that may impact outcomes, like delayed onset of cooling > 6 hours, due to the inherent limitations of the GHD.

A major limitation of the current study is that the time period of birth until referral and initiation of TH could not be extracted, as re-identification of babies that contribute more than one hospitalization is impossible because of pseudonymization and is strictly prohibited in the GHD. The treating center’s status was empirically derived from the dataset. Further, the origin of transfer (maternity hospital or lower-level pediatric hospital) and transferring center’s experience with BA could not be determined. Thus, the outborn status in this study implies that the infant was born in a center not adequately equipped to treat either the given severity of BA or its complications. Because no follow-up data are available in the GHD, only short-term outcomes could be assessed. The PCCC was used for this but has the important limitation that congenital malformations are extensively considered in the score. The relative proportions of congenital malformations were similar between both groups, making congenital malformations unlikely to be drivers of the observed differences in PCCC scores.

In spite of its limitations, the present study demonstrates clear differences in outcomes between inborns and outborns and has important implications for clinical practice. Infants who were postnatally transferred, indicating that they were born outside of a hospital prepared for their severity of disease, had higher risks for death and worse outcomes. Even if a cooling infrastructure is made widely available, the treatment of organ dysfunction in severely asphyxiated neonates requires a high level of experience that cannot be provided with comprehensive coverage. Thus, geographic coverage of cooling facilities must be provided by balancing the distribution of low-level cooling centers for moderate cases and high-level cooling centers equipped to treat organ dysfunction and provide organ replacement therapies. In 2023, the German Society for Perinatal Medicine demanded that obstetric departments should be mandatorily accompanied by pediatric departments to ensure the presence of neonatal expertise [29]. Hospitals should be equipped with staff specifically trained for neonatal emergencies, alongside with constant training, education and support by neonatal centers. The adoption of at least passive, preferably active servo-controlled cooling by experienced neonatal transport teams may improve access to adequate treatment within the critical time window and subsequently outcomes. This requires improved collaboration between facilities and the development of clear alarm protocols for referral including video consulting with tertiary care centers.

In conclusion, this study provides evidence that neonates with birth asphyxia who are born in centers without immediate access to adequate treatment display higher lethality and poorer short-term outcomes. Although the effects were attenuated in the subset analysis of high-risk infants receiving therapeutic hypothermia, they were still evident. These findings highlight the importance of ensuring that appropriate levels of neonatal care and resources are available for the management of birth asphyxia. Maternity hospitals in mandatory conjunction with pediatric departments would be ideal to improve outcomes of this highly vulnerable patient group.

## Data Availability

Data produced in this study cannot be provided upon request because the original data have to remain at the research data center. Any qualified researcher can file a request to access the data set at the site.

## Authors’ Contributions

Study design: NB, NF, UFM; preparation and planning of statistical analyses: NB, RH, AS, BK; statistical analyses: NB; data interpretation: NB, NF, UFM; drafting of the initial manuscript: NB, NF, UFM; critical revision of the results and manuscript: aSta, aSte, SK, CDS, RR.

## Conflicts of Interest and Financial Disclosures

The authors declare that they do not have conflicts of interest, including relevant financial interests, activities, relationships, and affiliations.

## Funding/Support and Role of Funder/Sponsor

The study received funding from the Stiftung Universitätsmedizin Essen.

## Data Access, Responsibility, and Anal

NB had full access to all the data in the study and takes responsibility for the integrity of the data and the accuracy of the data analysis.

## Data sharing statement

The original dataset remains at the FSO and can be accessed by qualified researchers at designated research data centers after filing a request and signing a confidentiality agreement.

---

**Figure S1:**
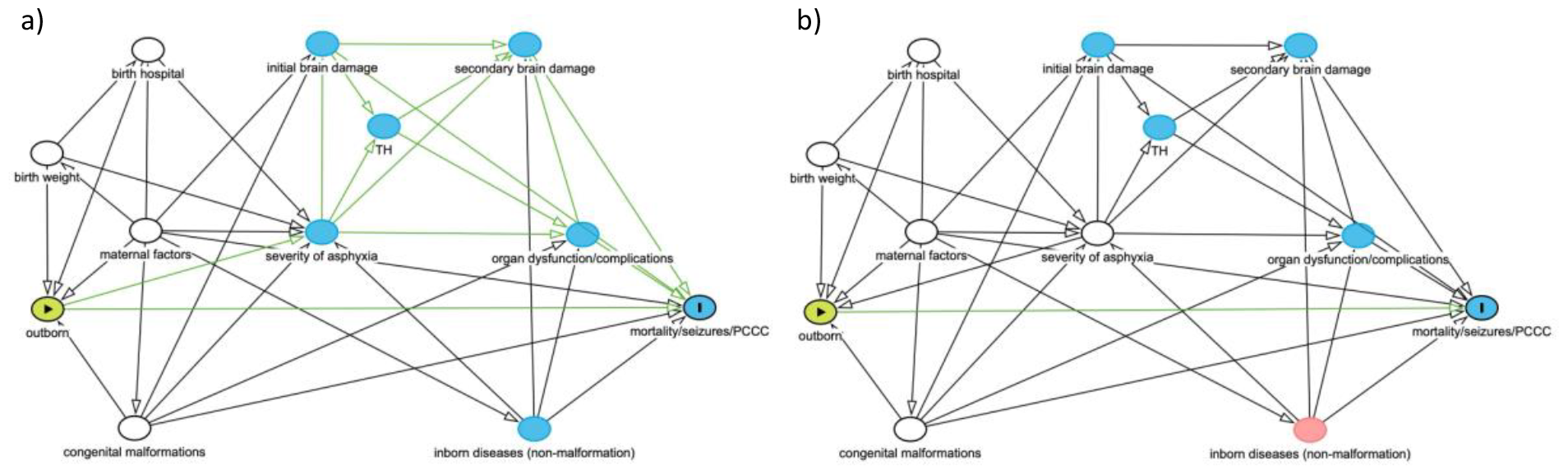
Directed acyclic graph to identify the minimally sufficient adjustment set for multivariable regression in neonates with birth asphyxia. a) Causal assumption: The birth hospital (and thus associated inborn/outborn status) determines the severity of asphyxia. The basic adjustment includes birth weight (in grams), maternal risk factors, and congenital malformations. The discharging hospital was accounted for as a random effect. b) Causal assumption: The severity of asphyxia determines the inborn/outborn status because severe asphyxia prompts transfer to a higher level center. The alternative adjustment set includes the severity of asphyxia as additional covariate. Green circle: exposure; blue circle with “I”: outcome; white circles: covariates in regression model.

## Notes

### Competing Interest Statement

The authors have declared no competing interest.

### Funding Statement

This study was funded by the Stiftung Universitaetsmedizin Essen

### Author Declarations

An existing routine data set from the Federal Statics Office of Germany available at the research data center Duesseldorf was used. No ethics approvement was necessary according to German law. All results were checked by the Federal Data Protection Officer.

