## Supplemental tables for "Short-term outcomes of asphyxiated neonates depending on outborn versus inborn status"

**Supplementary Table 1:** Extracted ICD- and OPS-Codes and newly calculated variables

| Diagnoses | ICD-10-Code(s) |
|---|---|
| Birth asphyxia | P21 (primary or secondary diagnosis) |
| Mild to moderate birth asphyxia | P21.1, P21.9 |
| Severe birth asphyxia | P21.0 |
| Birth weight | |
| Small for GA | P05, P07 |
| Large for GA | P08 |
| Maternal conditions | |
| Diabetes | P70.0, P70.1 |
| Maternal conditions unrelated to present pregnancy | P00 |
| Maternal complications of pregnancy | P01 |
| Complications of placenta, cord and membranes | P02 |
| Other complications of labor and delivery | P03 |
| Noxious substances transmitted via placenta or breast milk | P04 |
| Birth injuries | |
| Central nervous system | P10, P11.1, P11.2, P11.3, P11.4 |
| Peripheral nervous system | P14 |
| Skeletal | P13 |
| Other | P15 |
| Organ failure | |
| Cardiovascular (NOF_kard) | R57, P29 |
| Pulmonary (NOF_pulm) | P28, P22.0, P25, P27, |
| Coagulation (NOF_coag) | P60, P61.0 |
| Renal (NOF_ren) | N17 |
| Hepatic (NOF_hep) | K72 |
| Neurologic (NOF_CNS) | xxx |
| Neurologic complications | |
| Brain edema | P11.0 |
| Coma | P91.5 |
| Seizures | P90, G40 |
| Epileptic state | G41 |
| Hypoxic ischemic encephalopathy | P91.6 |
| Hydrocephalus | P91.7 |
| Congenital malformations | |
| Nervous system | Q00, Q01, Q02, Q03, Q04, Q05, Q06, Q07 |
| Eye, ear, face and neck | Q10, Q11, Q12, Q13, Q14, Q15, Q16, Q17, Q18 |
| Circulatory system | Q20, Q21, Q22, Q23, Q24, Q25, Q26, Q27, Q28 |
| Respiratory system | Q30, Q31, Q32, Q33, Q34 |
| Cleft lip and cleft palate | Q35, Q36, Q37 |
| Digestive system | Q38, Q39, Q40, Q41, Q42, Q43, Q44, Q45 |
| Genital organs | Q50, Q51, Q52, Q53, Q54, Q55, Q56 |
| Urinary system | Q60, Q61, Q62, Q63, Q64 |
| Musculoskeletal system | Q65, Q66, Q67, Q68, Q69, Q70, Q71, Q72, Q73, Q74, Q75, Q76, Q77, Q78, Q79 |
| Other | Q80, Q81, Q82, Q83, Q84, Q85, Q86, Q87, Q88, Q89 |
| Chromosomal abnormalities, not elsewhere classified | Q90, Q91, Q92, Q93, Q94, Q95, Q96, Q97, Q98, Q99 |

| **Procedures** | **OPS-Codes** |
|---|---|
| Dialysis | 8-854, 8-857, 8-858 |
| ECMO (extracorporeal membrane oxygenation) | 8-852 |
| Therapeutic hypothermia | 8-607 |
| **Other variables extracted from the basic data set** | |
| Sex | Included in the DRG data set |
| Length of stay (days) | Included in the DRG data set |
| Ventilation (hours) | Included in the DRG data set |
| Death | Reason for discharge = 7 |
| Early transfer | Reason for discharge = 6 or 8 and duration of hospital stay < 24 hours |
| Late transfer | Reason for discharge = 6 or 8 and duration of hospital stay≥ 24 hours |
| Early referral | Reason for admission = A (referral from external hospital within 24 hours and treatment duration in external hospital < 24 hours) |
| Birth | Reason for admission = G (birth) |
| Pediatric department | Treating department contains pediatric department code (10xx, 11xx, 12xx) |
| **Newly calculated variables** | **Variables/Procedures included [categories]** |
| Ventilation (dichotomous) | Ventilation (hours) not missing [yes, no] |
| Treatment center | [levels 1 - 3] |
| Obstetrics only | No case (any age < 18 years) treated in a pediatric department during study period [level 1] |
| Children's hospital without TH | Cases (any age < 18 years) treated in pediatric department but NO neonatal cases (age < 29 days) treated with hypothermia during study period [level 2] |
| Children's hospital with TH | Cases (any age < 18 years) treated in pediatric department AND neonatal cases (age < 29 days) treated with hypothermia during study period [level 3] |
| Neonatal organ failure score | Sum of binary scores NOF_kard, NOF_pulm, NOF_ren, NOF_coag, NOF_GI, NOF_hep, NOF_CNS, Dialysis, ECMO [continuous] |

**Supplementary table 2:** Calculation of Pediatric complex chronic conditions classification

| PCCC subscore | **ICD-10-Code(s)** |
|---|---|
| Neurological (CCC_neuro) | Q00, Q01, Q02, Q03, Q04, Q05, Q06, Q07, G901, F71, F72, F73, E75, F842, G11, G12, G25.3, G31.1, G31.8, G31.9, G32.8, G91.1, G93.8, G93.9, G94, G95.18, G95.88, G90.9, Q85.1, G80, G40, G37.1, G37.2, G37.8, G81, G82, G83.5, G83.9, G93.1, G93.5, R40, I63, G71, G72, G10, G20, G21.1, G21.8, G23.0, G23.1, G23.2, G24.0, G24.8, G25.3, G25.4, G25.5, G25.8, G25.9, T85.0, T85.1, T85.7, Z98.2, Z99 |
| Cardiological (CCC_cardio) | Q20, Q21.2, Q21.3, Q21.4, Q21.8, Q21.9, Q22, Q23, Q24, Q25.1, Q25.2, Q25.3, Q25.5, Q25.6, Q25.7, Q25.8, Q25.9, Q26, Q28.2, Q28.3, Q28.9, I34.0, I34.8, I36.0, I36.8, I37.0, I37.8, I42, I43, I51.5, I44, I45, I47, I48, I49, R00.1, I27, I50.9, I51.7, I51.8, I63.1, I63.2, Z95.1, T82.5, T82.1, T82.0, T82.2, T82.6, T82.7, Z95.0, Z95.2, Z95.3, Z95.8, Z95.9, Z45.01, Z45.02, Z45.09, T86.2, Z94.1 |
| Respiratory (CCC_resp) | Q30, Q31, Q32, Q33, Q34, P28.0, G47.32, I27.8, I43, J84.1, J96, Z90.2, E84, J95.0, J95.8, Z43.0, Z93.0, Z99.0, Z99.1, T86.81, Z94.2 |
| Renal (CCC_renal) | Q60, Q61, Q62, Q63, Q64, N18, Z90.5, Z90.6, G83.4, N31.2, N31.9, T85.71, Z93.5, Z93.6, Z91.1, Z99.2, Z43.5, Z43.6, Z46.6, T86.1, Z94.0 |
| Gastroenterological (CCC_gastro) | Q39.0, Q39.1, Q39.2, Q39.3, Q39.4, Q41, Q42, Q43, Q45, K73, K74, K75, K76.0, K76.1, K76.2, K76.3, K76.5, K76.8, K50, K51, I82.0, K55.1, K56.2, K59.3, Z98.0, Z90.3, Z90.4, Z93.1, Z93.2, Z93.3, Z93.4, Z43.1, Z43.2, Z43.3, Z43.4, T86.4, T86.8, Z94.4, Z94.88 |
| Hematological/immunological (CCC_hema_immu) | D55, D56, D57, D58, D60, D61, D71, D8, D72.0, M30.3, M35.9, D66, D68.2, D69.4, D69.3, D70.0, D70.5, D76.1, D76.2, D76.3, D86.9, B20, B21, B22, B23, B24, M30.0, M31.0, M31.3, M31.4, M31.6, M32.1, M33.9, M34.0, M43.1, M34.9 |
| Metabolic (CCC_metab) | E70.0, E70.2, E70.3, E70.8, E71, E72.0, E72.1, E72.3, E72.4, E72.8, E72.9, E74, E75, E77.0, E77.1, E78, E88.8, E76.0, E76.1, E76.2, E76.3, E85, E78.6, E79.1, E79.8, E80.4, E80.5, E80.6, E80.7, E93.0, E83.1, E83.3, E83.4, D84.1, E88, H49.8, E00.9, E23.0, E23.2, E23.3, E23.7, E24.0, E24.2, E24.3, E24.8, E24.9, E26.8, E25.0, E25.8, E25.9, Z46.8, Z96.4 |
| Genetic malformations (CCC_genet_malform) | Q90.9, Q91.3, Q91.4, Q91.7, Q92.8, Q93, Q95.0, Q96.9, Q97, Q98, Q99.8, Q99.9, E34.3, M41.0, M41.2, M41.39, M41.8, M41.9, M43.3, M96.5, Q72.2, Q75.0, Q75.2, Q75.9, Q76.0, Q76.1, Q76.2, Q76.4, Q76.5, Q76.6, Q77, Q78.0, Q87.1, Q78.2, Q78.3, Q78.4, Q78.8, Q78.9, K44.9, Q79.0, Q79.1, Q79.2, Q79.3, Q79.4, Q79.5, Q79.9, Q81, Q87.1, Q87.2, Q87.3, Q87.4, Q87.8, Q89.7, Q89.9, Q99.2 |
| Malignancy (CCC_malign) | C, D0, E34.0, D37, D38, D39, D4, Q85.0, T86.0, Z94.80, Z84.81 |
| Neonatal (CCC_neo) | P05, P07.0, P07.2, P10.0, P10.1, P10.4, P52.4, P52.8, P11.5, P21.0, P21.9, P96, P25, P27.0, P27.1, P27.8, P91.6, P35.0, P35.1, P35.2, P56.0, P57.0, P57.8, P61.3, P61.4, P77, P83.2, P91.2 |
| Dependency from technical devices (CCC_tech_dep) | T84.01, T84.02, T84.03, T84.04, T84.05, T84.06, T84.08, T84.11, T84.12, T84.18, T84.4, T84.5, T84.6, T84.7, T86.9, T86.0, T86.8, T86.5, T87.0, T87.1, Z99.8, Z93.1, Z93.2, Z93.3, Z93.4, Z43.1, Z43.2, Z43.3, Z43.4, T85.71, Z93.5, Z93.6, Z91.1, Z99.2, Z43.5, Z43.6, Z46.6, J95.0, J95.8, Z43.0, Z93.0, Z99.0, Z99.1, T82.5, |

| | |
|---|---|
| Transplantation recipient* (CCC_transplant) | T82.1, T82.0, T82.2, T82.6, T82.7, Z95.0, Z95.2, Z95.3, Z95.8, Z95.9, Z45.01, Z45.02, Z45.09, T85.0, T85.1, T85.7, Z98.2, Z99 Z94, T86 |
| PCCC score | Sum of binary sores CCC_neuro, CCC_cardio, CCC_resp, CCC_renal, CCC_gastro, CCC_hema_immu, CCC_metab, CCC_genet_malform, CCC_malign, CCC_neo, CCC_tech_dep, CCC_transplant |

* solid organ or bone marrow transplant
